# Serum protein profiling reveals a landscape of inflammation and immune signaling in early-stage COVID-19 infection

**DOI:** 10.1101/2020.05.08.20095836

**Authors:** Xin Hou, Xiaomei Zhang, Xian Wu, Minya Lu, Dan Wang, Meng Xu, Hongye Wang, Te Liang, Jiayu Dai, Hu Duan, Yingchun Xu, Xiaobo Yu, Yongzhe Li

**Affiliations:** Department of Clinical Laboratory & Beijing Key Laboratory for Mechanisms Research and Precision Diagnosis of Invasive Fungal Diseases, Peking Union Medical College Hospital, Chinese Academy of Medical Science & Peking Union Medical College, Beijing 100730, China; State Key Laboratory of Proteomics, Beijing Proteome Research Center, National Center for Protein Sciences-Beijing (PHOENIX Center), Beijing Institute of Lifeomics, Beijing, 102206, China

**Author notes:** These authors contributed equally to this work (Xin Hou, Xiaomei Zhang, Xian Wu, Minya Lu).

**Keywords:** SARS-CoV-2, COVID-19, proteomics, serum, biomarker, antibody microarray

## Abstract

Coronavirus disease 2019 (COVID-19) is a highly contagious infection and threating the human lives in the world. The elevation of cytokines in blood is crucial to induce cytokine storm and immunosuppression in the transition of severity in COVID-19 patients. However, the comprehensive changes of serum proteins in COVID-19 patients throughout the SARS-CoV-2 infection is unknown. In this work, we developed a high-density antibody microarray and performed an in-depth proteomics analysis of serum samples collected from early COVID-19 (n=15) and influenza (n=13) patients. We identified a large set of differentially expressed proteins (n=125) that participate in a landscape of inflammation and immune signaling related to the SARS-CoV-2 infection. Furthermore, the significant correlations of neutrophil and lymphocyte with the CCL2 and CXCL10 mediated cytokine signaling pathways was identified. These information are valuable for the understanding of COVID-19 pathogenesis, identification of biomarkers and development of the optimal anti-inflammation therapy.

## Introduction

The corona virus disease 2019 (COVID-19) is caused by the severe acute respiratory syndrome coronavirus 2 (SARS-CoV-2) and is highly contagious (Wang et al., 2020; Zhu et al., 2020). As of May 04, 2020, 3,524,429 cases of infection and 247,838 COVID-19 related deaths have been reported worldwide. The most common symptoms of COVID-19 are fever, malaise, fatigue, dry cough, conjunctivitis, diarrhea, and pneumonia (Huang et al., 2020a). While 19% of infected patients develop severe pneumonia or multiple organ failure, 81% have only mild symptoms or are asymptomatic. Individuals who are asymptomatic are a major challenge in controlling the COVID-19 pandemic since they are still contagious (Wu and McGoogan, 2020).

Recent studies have reported that COVID-19 patients with severe symptoms have elevated plasma levels of IL2, IL6, IL7, IL10, GCSF, IP10, MCP1, MIP1A, and TNFα (Chen et al., 2020b; Huang et al., 2020a; Mehta et al., 2020; Ruan et al., 2020), which suggests that the progression of COVID-19 is associated with cytokine storms and immunosuppression. Further profiling of the proteins in serum may help to understand the pathogenesis of COVID-19 and to develop effective treatment (Huang et al., 2020b; Zhang et al., 2020). However, to date, there is neither a vaccine to prevent COVID-19 nor an effective medicine to treat COVID-19.

In this study, a comprehensive analysis of serological proteins from patients in the early stages of COVID-19 or influenza was performed using an antibody microarray developed in our laboratory (Xu et al., 2019). A large number of differentially-expressed proteins were identified that mediates diverse immune signaling pathways related to viral infection, the inflammatory response, and immune cell activation and migration. These data reflect the host response to COVID-19 infection, thus providing insight into COVID-19 pathogenesis and providing potential therapeutic targets.

## Methods

### Samples

All COVID-19 and influenza patients were enrolled in the outpatient department of the Peking Union Medical College Hospital (Peking, China). Serum samples were collected with written informed consent under the approval of the intuitional review board (IRB) from the Peking Union Medical College Hospital (Ethical number: ZS-2303) and the Beijing Proteome Research Center (Beijing, China). All experiments were performed according to the standards of the Declaration of Helsinki. COVID-19 patients with mild symptoms were positively diagnosed using reverse transcription PCR (RT-PCR) that detects SARS-CoV-2 RNA. The testing was in accordance to the Diagnosis and Management Plan of Pneumonia with Novel Coronavirus Infection (trial version 7).

### Screening of COVID-19 serum proteome using antibody microarrays

The antibody microarrays and sera screening were processed as previously described (Xu et al., 2019). Briefly, 10 μL of serum were diluted with 90 μL phosphate buffered saline (PBS; pH 7.4) and labeled with 1 μL of NHS-PEG4-Biotin (20 g/L in DMSO) (Thermo Fisher Scientific, MA, USA) for 1 h at room temperature. After removing the excess biotin molecules, the biotinylated proteins were dissolved in 400 μL of PBS containing 5% milk (w/v). In parallel with biotin labeling, antibody microarrays were blocked with 500 μL of PBS with 5% milk (w/v) for 1 h at room temperature. After removing the milk, the arrays were incubated with biotin-labeled proteins at 4 °C overnight. The slides were washed with PBS containing 0.05% (w/v) Tween 20 (PBST). To detect the bound proteins, the arrays were incubated with 2 μg/mL streptavidin-Phycoerythrin (PE) (Jackson Immunoresearch, USA) for 1 h at room temperature in the dark and then washed with PBST. After centrifuging for 2 min at 1000×g, the slide was scanned using the GenePix 4300A microarray scanner at the wavelength of 532 nm.

### Bioinformatics analysis

Functional annotation of protein classes was performed using the PANTHER database (http://pantherdb.org/)(Mi et al., 2016). The GO biological process analysis was performed using CluGO and visualized in Cytoscape (version 3.7.2) using two-sided hypergeometric test with a p-value less than 0.01 (Bindea et al., 2009; Franz et al., 2016). The analyses of signaling pathways, protein domains and cellular components were performed using the STRING database (Szklarczyk et al., 2015).

### Statistical analysis

Differentially-expressed proteins were identified using the t-distributed t-test (p-value = 0.05). Hierarchical clustering was performed using the Mev software (Chu et al., 2008; Yu et al., 2013).

## Results

### In-depth profiling of the serum proteome in early-stage COVID-19 and influenza patients

The COVID-19 status of twenty-eight patients with similar symptoms (i.e. fever, headache, cough sputum, myalgia) were ascertained via RT-PCR, which detects SARS-CoV-2 RNA. In addition, the patients were tested 2 – 4 days from symptom onset. Fifteen (15) patients were positive for COVID-19 while 13 patients were negative for COVID-19; this latter group was classified as influenza (Figure 1A, Table 1). The proteins in the serum were measured using an antibody microarray containing 532 antibodies developed in-house and as previously described (Supplementary Table 1, Figure 1B) (Xu et al., 2019). The antibodies on the array target intercellular signaling molecules, protein-binding activity modulators, protein modifying enzymes and metabolite interconversion enzymes (Figure 1C). The intra- and inter-reproducibility of the microarray are 0.9976 and 0.999, respectively (Figure1D).

**Figure 1.**
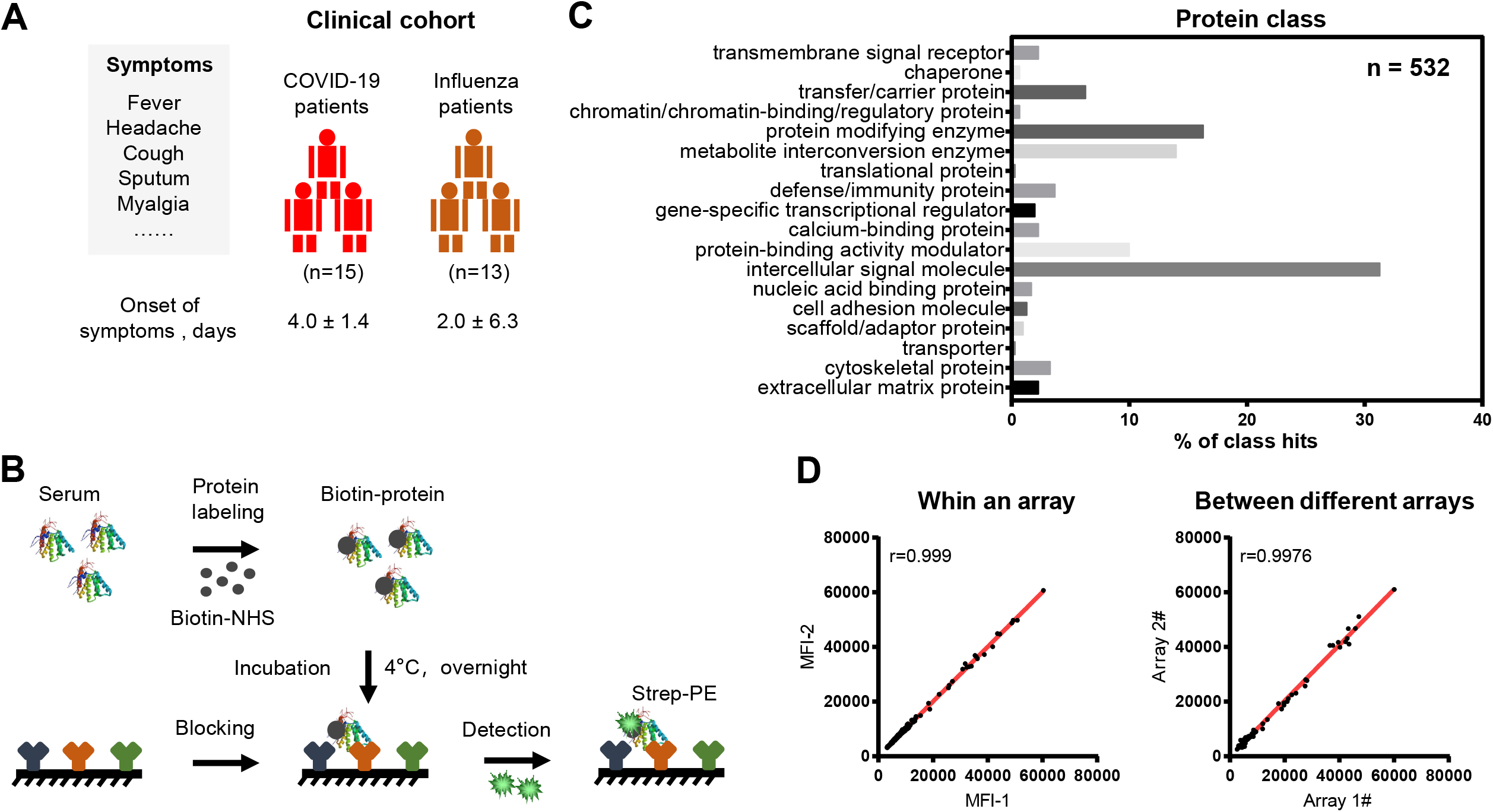
Schematic illustration of serum screening of early-stage COVID-19 patients. (A) Characteristics of the clinical cohort used in this study. (B) Workflow for serum screening using the antibody microarray; (C) Classes of proteins on the antibody microarray; (D) Reproducibility of the antibody microarray for the detection of serum proteins.

**Table 1.** Characteristics of early-stage COVID-19 patients.

|  | **Influenza patients**<br>**(n=13)** | **COVID-19 patients**<br>**(n=15)** |
| --- | --- | --- |
| **Male** | 8 (61.5%) | 7 (46.7%) |
| **Female** | 5 (38.5%) | 8 (53.3%) |
| **Age, year** | 51.0 (20.0-88.0) | 38.0 (7.0-68.0) |
| **SARS-CoV-2 (+)** | 0 | 15 (100%) |
| **FluA-RNA(+)** | 6 (46.2%) | 0 |
| **FluB RNA(+)** | 3 (23%) | 0 |
| **RSV RNA(+)** | 4 (30.8%) | 0 |
| **Exposure history** | 4 (30.8%) | 14 (93.3%) |
| **Fever** | 13 (100%) | 11 (73.3%) |
| **Headache** | 4 (30.8%) | 1 (6.7%) |
| **Cough** | 6 (46.2%) | 7 (46.7%) |
| **Sputum** | 3 (23%) | 2 (13.3%) |
| **Myalgia** | 7 (53.8%) | 2 (13.3%) |
| **Fatigue** | 6 (46.2%) | 0 |
| **Diarrhea** | 1 (7.7%) | 0 |
| **Dyspnea** | 1 (7.7%) | 2 (13.3%) |
| **Nausea or vomiting** | 0 | 0 |
| **Onset of symptom ,**<br>**Median ± SD, days** | 2.0 ± 6.3 | 4.0 ± 1.4 |
| **Pulmonary ground-glass**<br>**opacity** | 7(53.8%) | 12 (80%) |
| **Bilateral pulmonary**<br>**infiltration** | 6(46.2%) | 10 (66.7%) |

Data are median (IQR) or n (%).

Using a t-distributed t-test, 88 up-regulated and 37 down-regulated proteins between COVID-19 and influenza group patients were identified (p-value < 0.05) (Figure 2A and 2B, Supplementary Table 2). Among of them, the elevation of five cytokines (IFNG, IL6, CXCL8, CXCL10 and CCL2) were reported in mild and severe COVID-19 patients (Figure 2C) (Huang et al., 2020b; Schett et al., 2020; Wu and Yang, 2020). These results demonstrate that the microarray is reliable in detecting serum proteins from COVID-19 patients. Many differentially-expressed proteins not reported to be associated with COVID-19 infection were also detected, including IL20, CCL27, IL21, PLG, C1R and C7 (Figure 2D).

**Figure 2.**
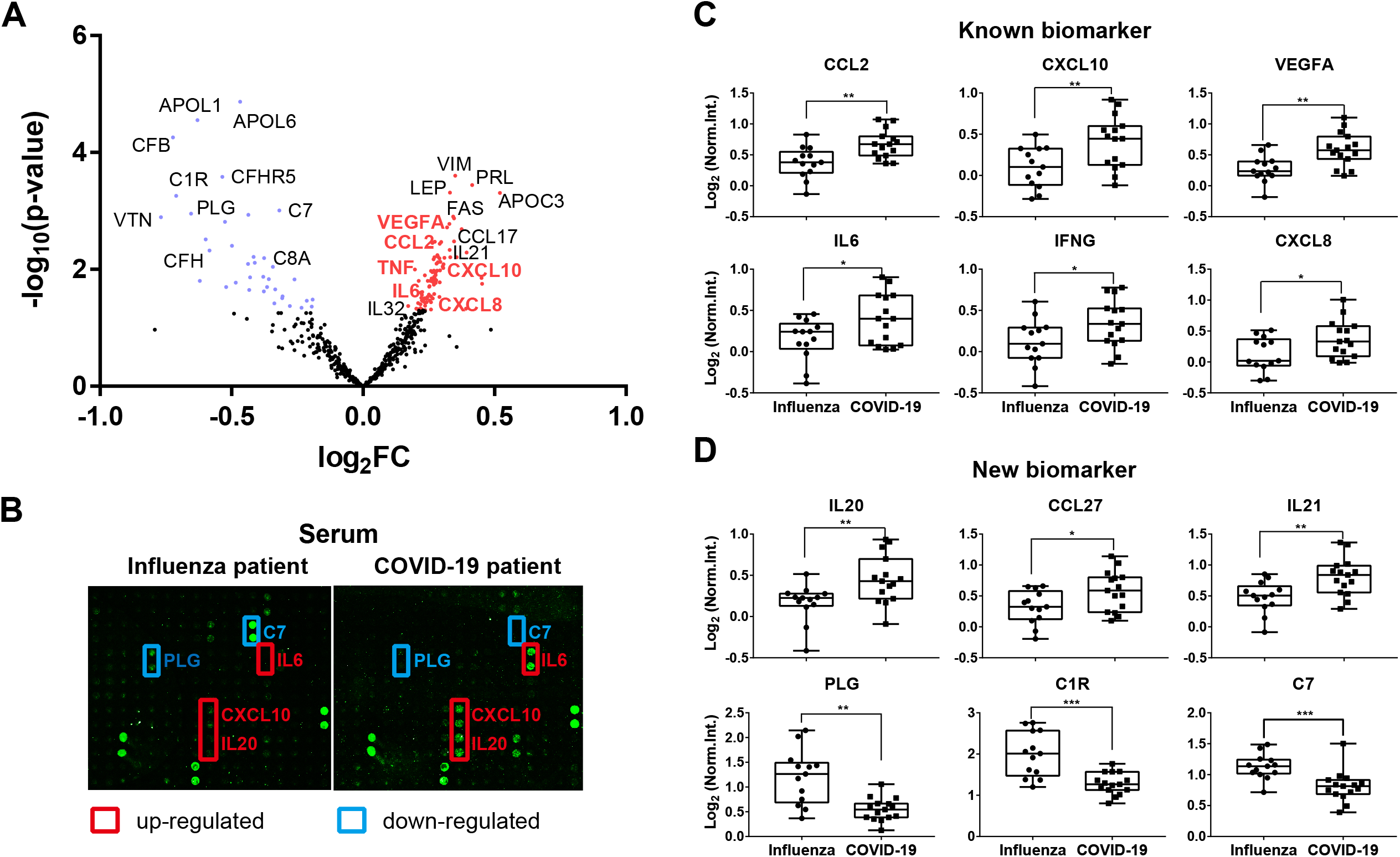
Detection of proteins associated with early-stage COVID-19. (A) Volcano plot analysis of COVID-19 associated proteins in serum. Blue and red dots represent down-and up-regulated proteins in early-stage COVID-19, respectively. Protein symbols in red represent the serum proteins associated with COVID-19 reported in the literature; (B) Representative results of antibody microarray images in the serum of early-stage COVID-19 and influenza patients; (C) Box plot analysis of known COVID-19 associated serum proteins; (D) Box plot analysis of newly identified COVID-19 associated serum proteins. *, ** and *** represent the p-value less than 0.05, 0.01 and 0.001, respectively.

All of the COVID-19 patients were distinguished from influenza patients using hierarchical cluster analysis of the differentially-expressed proteins (Figure 3A). These data suggest that these proteins could be potential biomarkers for early COVID-19 diagnosis. A large independent cohort should be used to validate these findings.

**Figure 3.**
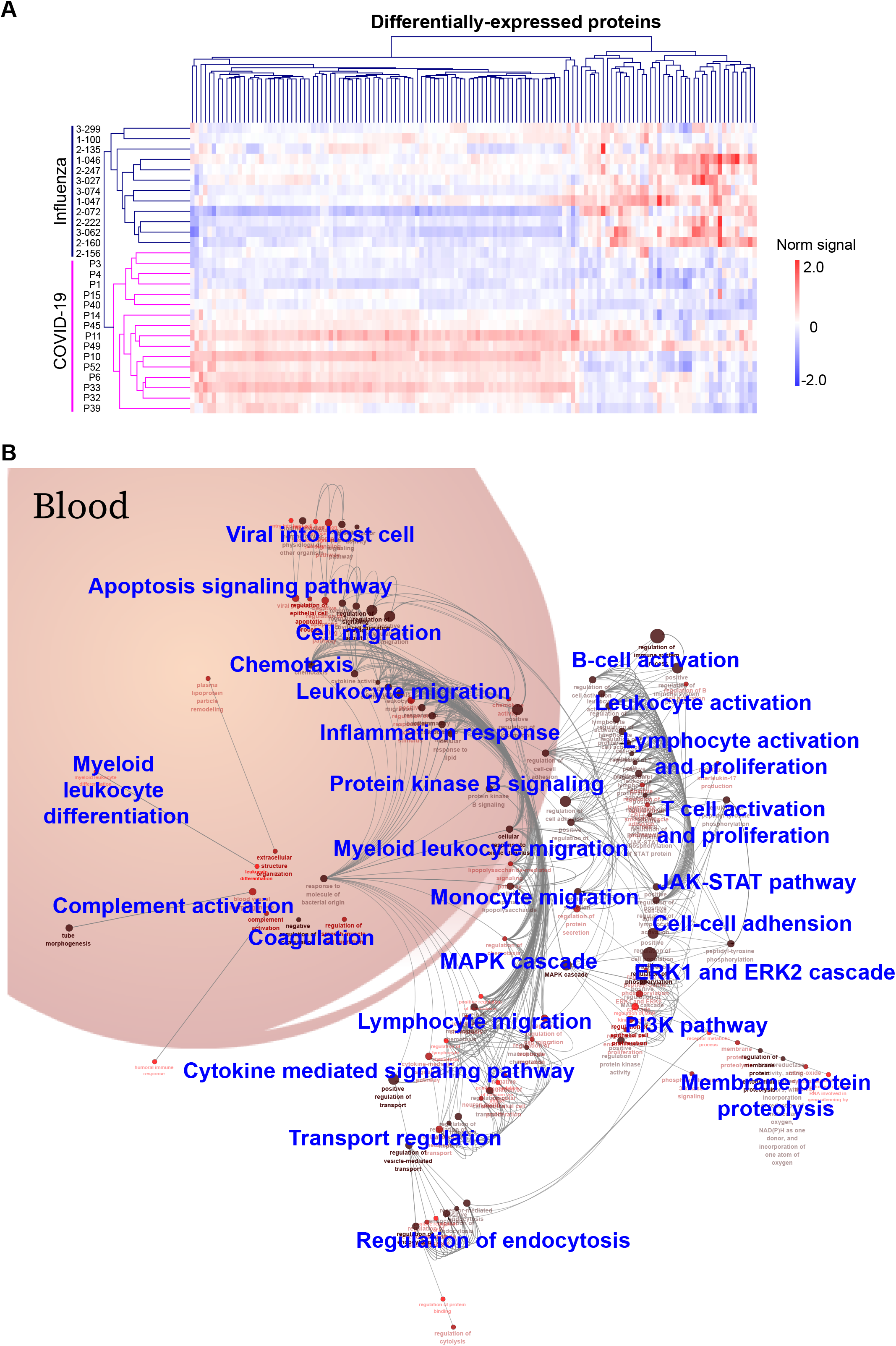
A landscape of immune signaling in early SARS-CoV-2 infection. (A) Hierarchical clustering analysis of differentially-expressed proteins between the COVID-19 and influenza patient groups with a p-value < 0.05 (t-distributed t test). Intensities of proteins were log2-transformed. (B) GO biological process analysis visualized in Cytoscape. Significantly enriched biological processes were selected using a two-sided hypergeometric test with a p-value < 0.01.

### Prevalent inflammation and immune signaling in early COVID-19 infection

Twenty-six biological processes were identified from GO analysis of the differentially-expressed proteins in early-stage COVID-19 (Figure 3B, Supplementary Table 3). The biological processes are divided into four classes: (1) immune cell activation and migration, (2) cellular activities, (3) protein signaling that regulate viral infection, and (4) blood functional systems. More specifically, the first class of biological processes includes T cells, B cells, monocytes, myeloid leukocytes, etc. The second class of biological processes mainly reflects cellular activities, such as endocytosis, membrane protein proteolysis, cell-cell adhesion. The third class of biological processes include signaling pathways that regulate the viral infection, including the MAPK, ERK1/ERK2, JAK-STAT, and PI3K pathways. The fourth class of biological processes is blood functional systems, including inflammation response, complement, coagulation and chemotaxis.

Next, the up- and down-regulated signaling pathways in early SARS-CoV-2 infection were further selected using STRING by filtering for differentially-expressed proteins (n≥10) and a p-value less than 0.01 (Figure 4A, Supplementary Table 4). Signaling pathways that were up-regulated the most were cytokine-cytokine receptor interaction, cytokine signaling in immune system, IL-4 and IL-13 signaling and IL-17 signaling. Signaling pathways representing the second most up-regulated group were associated with viral infection, including the JAK-STAT, MAPK, and PI3K-Akt signaling pathways. In contrast, neutrophil degranulation, complement cascade, and coagulation cascades were down-regulated in early COVID-19 infection.

**Figure 4.**
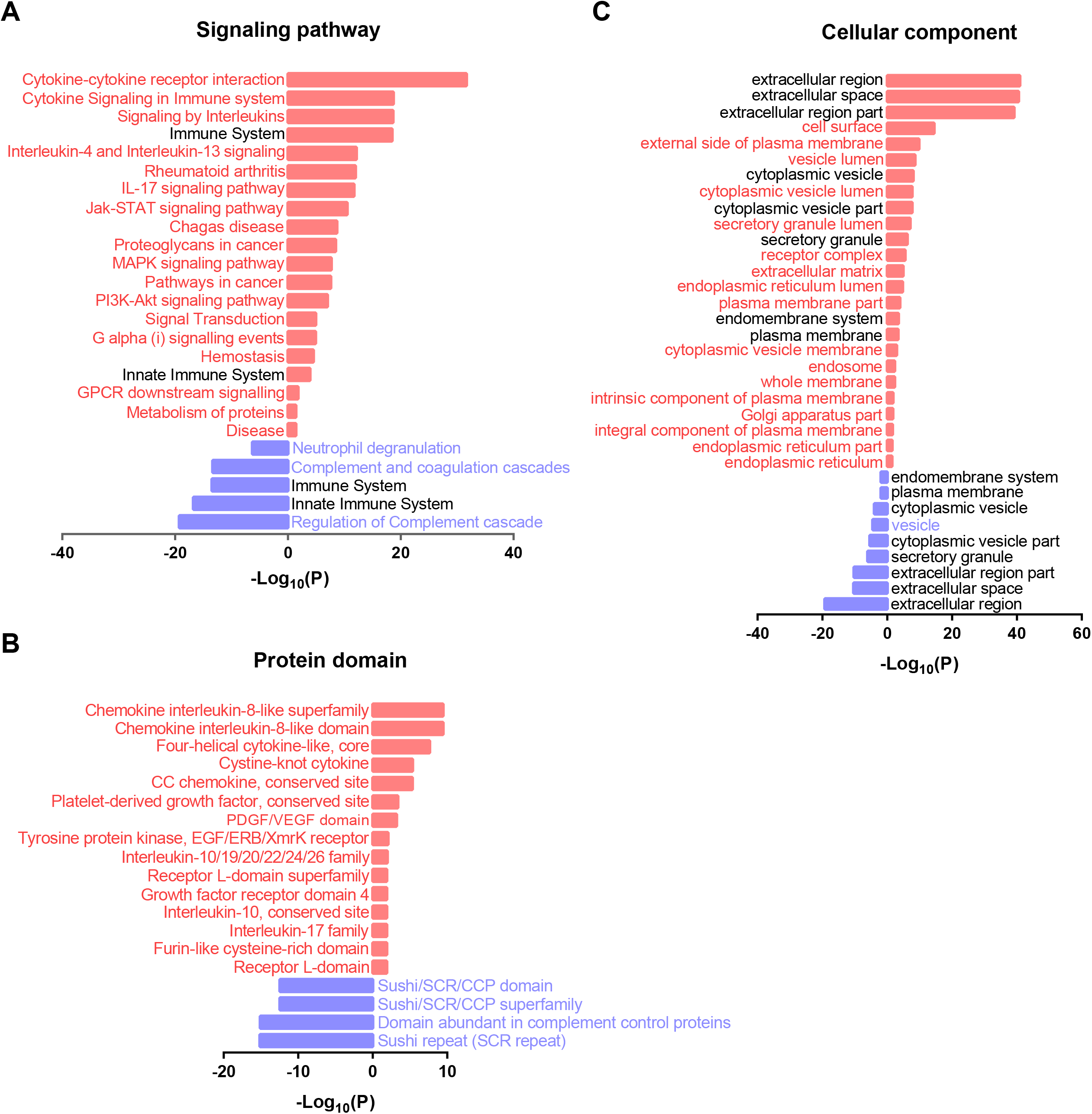
Functional annotation of the differentially-expressed proteins in early SARS-CoV-2 infection. (A), (B), and (C) are the significantly altered signaling pathway, protein domain, and cellular component identified by antibody microarrays in early-stage COVID-19, respectively. P-values were calculated by two-sided hypergeometric test based on the KEGG/Reactome/InterPro database and GO terms. Upregulated and downregulated pathways are indicated in red (right) and blue (left) bars.

The most conserved protein domains within the differentially-expressed proteins were also from the cytokine and chemokine protein family, including the cytokine interleukin-8 like superfamily, chemokine interleukin-8-like domain, four-helical cytokine-like, core, cysteine-knot cytokine, CC chemokine, conservation site, interleukin-10/19/20/22/24/26 family, interleukin-10 conservation site and interkin-17 family (Figure 4B). The second most abundant protein domains were from platelet-derived growth factor (PDGF) conservation site, PDGF/VEGF domain and growth factor receptor domain 4. In addition, the domains of tyrosine protein kinase (EGF/ERB/XmrK receptor), receptor L-domain superfamily and receptor L-domain were identified. By contrast, the domains of down-regulated proteins were the sushi/SCR/CCP domain, sushi/SCR/CCP family, and sushi repeat (SCR repeat).

Functional annotation of the cellular components shows that the up-regulated proteins were specifically enriched in cell surface, external side of plasma membrane, vesicle lumen, cytoplasmic vesicle lumen, receptor complex, extracellular matrix, endoplasmic reticulum lumen, plasma membrane part, cytoplasmic vesicle membrane, endosome, whole membrane, intrinsic component of plasma membrane, Golgi apparatus part, integral component of plasma membrane, endoplasmic reticulum part and endoplasmic reticulum. Only one cellular component was enriched in the down-regulated protein: the vesicle (Figure 4C).

To highlight the involvement of cytokines in COVID-19 infection, 29 chemokines, cytokines, or growth factors are involved in the cytokine-cytokine receptor and viral protein interaction with cytokine and cytokine receptor pathways (Figure 5). These include fifteen chemokines (BAFF, CX3CL1, CCL2, CCL4, CCL4L1, CCL4L2, CCL5, CCL8, CCL10, CCL12, CCL16, CCL17, CCL27, CCL28, CCL32), twelve cytokines (IFNG, IL3, IL6, IL12, IL13, IL17A, IL17B, IL17E, IL19, IL20, IL21, IL32) and two growth factors (TGFB1, NGF).

**Figure 5.**
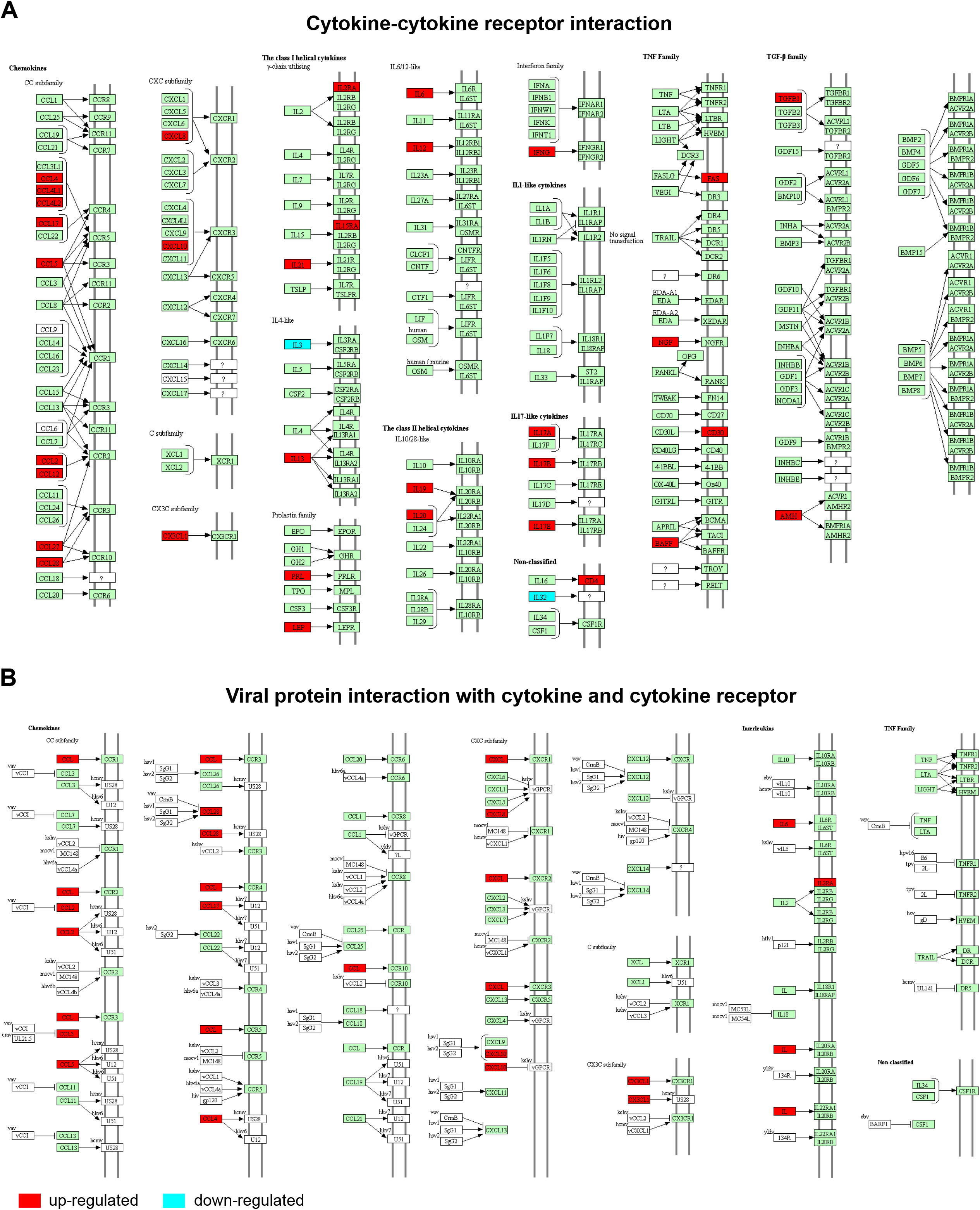
The cytokine mediated signaling pathways associated with early SARS-CoV-2 infection. (A) Cytokine-cytokine receptor interaction; (B) Viral protein interaction with cytokines and cytokine receptors. Red and blue boxes represent up- and down-regulated genes, respectively.

### CCL2 and CXCL10 medicated cytokine signaling pathways significantly correlate with neutrophil and lymphocyte respectively

A comprehensive correlation analysis of the differentially-expressed proteins and clinical indices in early-stage COVID-19 patients was performed (Figure 6, Supplementary Table 5). Protein expression was positively correlated with clinical indices reflecting the liver function (Albumin, Alb; Alanine aminotransferase, ALT; Total bilirubin, TBil), renal function (Creatinine, Cr(E); Urea), acute myocardial injury (Creatine Kinase MB mass, CKMB-mass; Cardiac Troponin I, cTnI; N-terminal pro-Brain Natriuretic Peptide, NT-proBNP; Creatine Kinase, CK), and inflammation and infection (C-reactive protein, CRP; White Blood Cell, WBC; Neutrophil, NEUT) (Figure 6A). By contrast, the expression of some proteins had a negative correction with the lymphocyte (LY) count, eosinophil percentage (EOS%) and Direct Bilirubin (DBil) level (Figure 6B).

**Figure 6.**
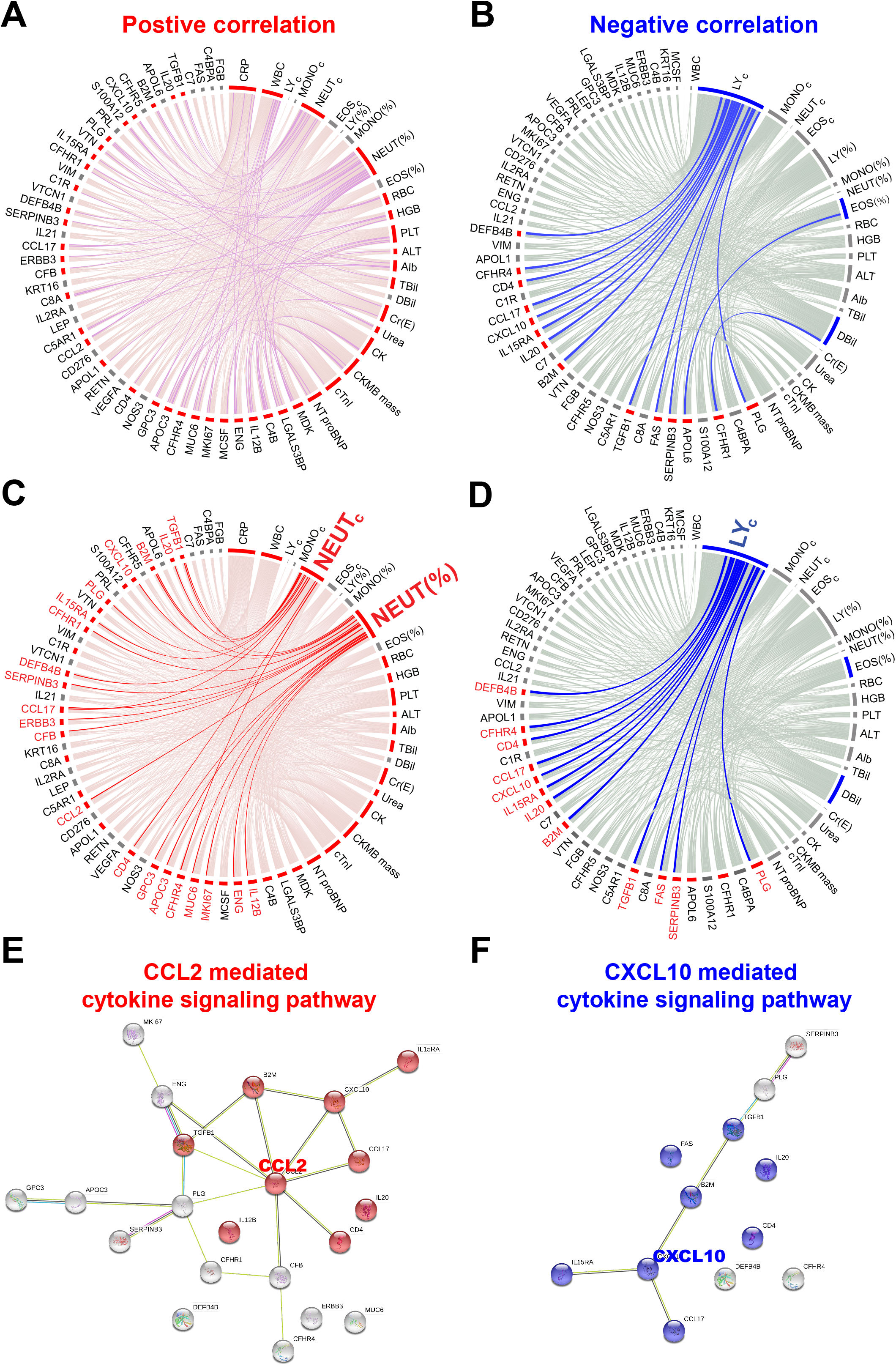
Correlation network of early COVID-19 specific proteins and clinical indices. (A) and (B) are the positive and negative correlations of early SARS-CoV-2 infections and twenty clinical indices using circus, respectively. Correlations with statistical significance (p-value < 0.05) are indicated in pink. Non-significant (p-value ≥ 0.05) positive and negative correlations are indicated in red and blue, respectively. (C) and (D) are proteins correlated with neutrophils and lymphocytes, respectively. (E) and (F) are the protein-protein interaction networks of COVID-19 proteins that are positively and negatively associated, respectively, with neutrophils and lymphocytes. The protein-protein interactions and KEGG pathway analysis were performed in the STRING database (https://string-db.org/).

Among these clinical indices, NEUT and LY have the largest correlations with 21 and 12 proteins in COVID-19 patients, respectively (Figure 6C and 6D). The functional annotation of these proteins using theSTRING database show that the proteins are enriched in the CCL2- and CXCL10-mediated cytokine signaling pathways, respectively (Figure 6E and 6F).

## Discussion

Understanding the host’s response to SARS-CoV-2 infection is crucial to designing a therapeutic regime to efficiently clear the SARS-CoV-2 virus and minimize tissue damage (Shi et al., 2020). An abundance of evidence has revealed that cytokines play an important role in the severity of COVID-19 symptoms (Chen et al., 2020a; Huang et al., 2020b; Shi et al., 2020). However, the overall protein expression changes in serum during early SARS-CoV-2 infection is unknown.

To address this concern, we fabricated a high-density antibody microarray to profile the expression of 532 serological proteins of early-stage COVID-19 and influenza patients (Figure 1). One primary advantage of antibody arrays compared to another popular protein detection tool, mass spectrometry, is that depletion of high abundance proteins is unnecessary and ion suppression is not a concern. Thus, low abundance proteins in serum can be identified with antibody arrays with minimal sample processing (Xu et al., 2019).

We found that differentially-expressed proteins (n=125 p-value < 0.05) (Figure 2) participate in a broad range of biological processes associated with viral infection, inflammation, immune cell activation and migration, and the complement and coagulation processes (Figure 3B). Changes in the complement and coagulation cascades have also been identified in early SARS-CoV-2 infection and severe COVID-19 patients (Campbell and Kahwash, 2020; Risitano et al., 2020; Tanaka et al., 2016; Tang et al., 2020). Our data reveal that activation of viral infection pathways (MAPK, ERK1/ERK2, JAK-STAT, PI3K) occurs in acute COVID-19 infection. Previous studies have shown that the MAPK signaling pathway is a major immune response to SARS-CoV-2 infection, and that the ERK and PI3K signaling pathways play a critical role in MERS-CoV and SARS-CoV pathogenesis (Huang et al., 2020c) (Kindrachuk et al., 2015; Mizutani et al., 2005).

We show that cytokine-mediated signaling pathways are the major class of dysregulated pathways in early SARS-CoV-2 infection (Figure 4A and 4B, Figure 5). These pathways also play integral roles in SARS-CoV-1 infection (Channappanavar et al., 2016). The expression changes of these cytokines (i.e., IL6, IL17, CCL2) indicate that pro-inflammation may occur in early stages of SARS-CoV-2 infection (Figure 1A, Table 1). In addition, our study identified extensive upregulation of chemokines, which are often stimulated by pro-inflammatory cytokines and mainly function as chemoattractants for effector cells (e.g., leukocytes, recruiting monocytes, neutrophils) from the blood to sites of infection or tissue damage.

The CCL2- and CXCL10-mediated cytokine signaling pathways, which are known to be involved in viral infections, were enriched in a subset of differentially-expressed proteins in early SARS-CoV-2 infection (Figure 6C-F). CCL2 is a CC chemokine that attracts monocytes, memory T lymphocytes, and basophils, and is associated with lung inflammatory disorders, including acute respiratory distress syndrome, asthma, and pulmonary fibrosis. Notably, these inflammatory disorders and pulmonary infiltration are associated with the progressive respiratory failure and death of SARS patients (Chen et al., 2010). Furthermore, CCL2 can act as an autocrine factor that promotes viral replication in infected macrophages, and CCL2 neutralization inhibits HIV-1 replication in monocyte-derived macrophages (Sabbatucci et al., 2015). Interestingly, a novel class of small molecule inhibitor, bindarit, alleviates inflammation caused by Chikungunya and Ross River viruses by reducing CCL2 synthesis in animal models (Rulli et al., 2009; Rulli et al., 2011). CXCL10 is a ligand for the CXCR3 receptor, the activation of which results in the recruitment of T cells and the perpetuation of mucosal inflammation. Tacrolimus (FK506), a macrolide immunosuppressive agent isolated from *Streptomyces tsukubaensis,* has been found to suppress CCL2 and CXCL10 expression in human colonic myofibroblasts (Aomatsu et al., 2012). All the results combined suggest that CCL2 and CXCL10 may have the potential to be used as anti-inflammation targets for COVID-19 therapy (Liu et al., 2011; Zhang et al., 2020).

There are three limitations in this study. First, the number of serum samples was limited; thus, the biomarkers identified in our study should be validated in a large independent clinical cohort. Second, protein detection depends on the sensitivity and specificity of the capture antibodies immobilized on the microarray. Third, serological proteins of early-stage COVID-19 patients were only compared to early-stage influenza patients. In the future, protein profiling of COVID-19 patients should be examined over the entire course of infection. In addition, the profiles should be compared to healthy patients and patients infected with different viruses.

Our study comprehensively profiled the serological proteins of early SARS-CoV-2, revealing a new understanding of the inflammation and immune signaling that occurs. Our data also identified potential biomarkers that could be used to diagnose COVID-19 patients and design effective treatment.

## Data Availability

The data is avaialable upon of requrest.

## Contributions

X. H., Y. L. and X. W. provided the clinical samples. X. Z., H. W., D. W., J. D., T. L. and M. X. prepared the microarrays. X. H., X. Z., and M. L. executed microarray experiments. X. Z., D. W., T. L.. H. D. and X.Y. executed the statistical and structural analysis. X.Y., Y. X. and Y. L. conceived the idea, designed experiments, analyzed the data and wrote the manuscript.

## Acknowledgement

This research was supported by grants from the National Natural Science Foundation of China Grants (81671618, 81871302, 81673040, 31870823), CAMS Innovation Fund for Medical Sciences (CIFMS) (2017-I2M-3-001, 2017-I2M-B&R-01), the State Key Laboratory of Proteomics (SKLP-C202001, SKLP-O201703, SKLP-K201505), the Beijing Municipal Education Commission and the National Program on Key Basic Research Project (2018YFA0507503, 2017YFC0906703, 2018ZX09733003). We also thank Dr. Brianne Petritis for her critical review and editing of this manuscript.

## Supplementary information

The supplementary information includes six supplementary tables.

